# Monitoring SARS-CoV-2 variants alterations in Nice neighborhoods by wastewater nanopore sequencing

**DOI:** 10.1101/2021.07.09.21257475

**Authors:** Géraldine Rios, Caroline Lacoux, Vianney Leclercq, Anna Diamant, Kévin Lebrigand, Adèle Lazuka, Emmanuel Soyeux, Sébastien Lacroix, Julien Fassy, Aurélie Couesnon, Richard Thiery, Bernard Mari, Christian Pradier, Rainer Waldmann, Pascal Barbry

## Abstract

**Background:** Wastewater surveillance has been proposed as an epidemiological tool to define the prevalence and evolution of the SARS-CoV-2 epidemics. However, most implemented SARS-CoV-2 wastewater surveillance projects were relying on qPCR measurement of virus titers and did not address the mutational spectrum of SARS-CoV-2 circulating in the population.

**Methods:** We have implemented a nanopore RNA sequencing monitoring system in the city of Nice (France, 550,000 inhabitants). Between October 2020 and March 2021, we monthly analyzed the SARS-CoV-2 variants in 113 wastewater samples collected in the main wastewater treatment plant and 20 neighborhoods.

**Findings:** We initially detected the lineages predominant in Europe at the end of 2020 (B.1.160, B.1.177, B.1.367, B.1.474, and B.1.221). In January, a localized emergence of a variant (Spike:A522S) of the B.1.1.7 lineage occurred in one neighborhood. It rapidly spread and became dominant all over the city. Other variants of concern (B.1.351, P.1) were also detected in some neighborhoods, but at low frequency. Comparison with individual clinical samples collected during the same week showed that wastewater sequencing correctly identified the same lineages as those found in COVID-19 patients.

**Interpretation:** Wastewater sequencing allowed to document the diversity of SARS-CoV-2 sequences within the different neighborhoods of the city of Nice. Our results illustrate how sequencing of sewage samples can be used to track pathogen sequence diversity in the current pandemics and in future infectious disease outbreaks.

## Introduction

New strategies are currently investigated to track the emergence and spread of SARS-CoV-2 variants. Epidemiological surveillance has largely relied on individual testing by quantitative real-time polymerase chain reaction (RT-qPCR) or digital droplet polymerase chain reaction (ddPCR). However, a patient-oriented approach is hardly sustainable for a long period in a large population. Individual testing has also some intrinsic limitations. Information is mostly derived from symptomatic cases and does not necessarily reflect the full dynamics of COVID-19 diffusion. The incomplete identification of asymptomatic, pre-symptomatic or post-symptomatic cases by individual testing can be overcome by analyses of pooled samples that reflect the entire population [1]. The principal sites of SARS-CoV-2 replication are the upper and lower respiratory tracts, but the virus also replicates in the intestine, leading to high concentrations of SARS-CoV-2 in human stool [2]. In consequence, analysis of SARS-CoV-2 in wastewater appears a promising approach for cost-effective population-scale SARS-CoV-2 epidemiology. Several recent studies that analyzed SARS-CoV-2 titers in Wastewater Treatment Plants (WWTPs) by qPCR revealed a good correlation between SARS-CoV-2 incidence rates and the virus titers in wastewater [3-8].

The emergence of variants of concern (VOCs) associated with higher transmissibility and/or immune evasion in England (B.1.1.7), South Africa (B.1.351), South America (P.1), the United States of America (B.1.525) and India (B.1.617.2) recently added a further layer of complexity. Measurement of virus concentrations needs now to be complemented by a direct quantification of the individual VOCs. This can be done with variant-specific RT-qPCR/ddPCR assays, but the continuing emergence of new variants calls for more generic approaches.

High throughput sequencing (HTS) of SARS-CoV-2 overcomes the limitations of qPCR since it allows the identification of both known and novel mutations in the virus sequence. HTS also allows the definition of the relative abundance of individual virus variants in a sample and thus is well suited for the analysis of complex virus populations in samples such as wastewater.

Our aim was to evaluate a wastewater SARS-CoV-2 sequencing approach in a real-life configuration. We performed this study in Nice (France). Samples covering 416,755 inhabitants were collected from the inlet of the wastewater treatment plant and upstream in sewers draining different neighborhoods once a month between October 2020 and March 2021. We show here that nanopore sequencing of wastewater not only revealed the dynamics of known SARS-CoV-2 lineages within the city of Nice but also allowed the identification of novel variants that would have escaped identification with variant-specific qPCR assays.

## Results and Discussion

Our goal was to analyze lineage composition in multiple neighborhoods, including specific areas that are more susceptible to outbreaks (*e*.*g*. zones with a lower standard of living, higher population density). We collected 113 distinct wastewater samples from 21 different locations spread all over the territory of Nice (Figure 1A-B, Supplementary Table 1) once a month between October 2020 and March 2021. SARS-CoV-2 cDNA was amplified using the tiling PCR approach developed by the ARTIC network [9] and sequenced on Oxford Nanopore sequencing devices (Figure 1C, methods section).

**Figure 1:**
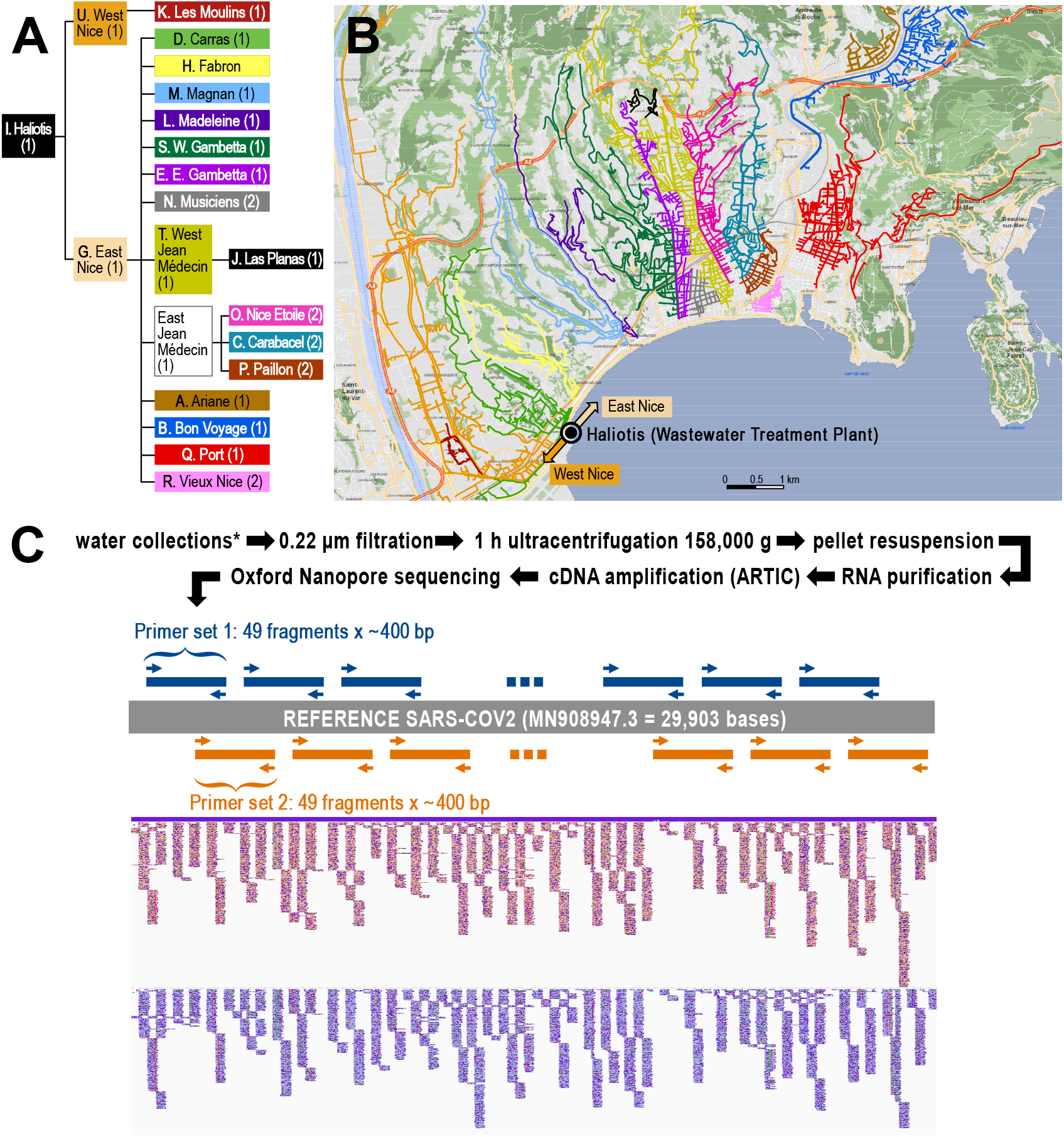
Experimental design of the study. **(A)** Hierarchical organization of the sampling points. **(B)** Map of the Nice area, with the indications of the different catchment areas. Haliotis is the name of the central wastewater treatment plant (WWTP). **(C)** flow-chart of the project, with the different steps of analysis. The ARTIC sequencing protocol is based on a polymerase chain amplification of 2 sets of non-overlapping amplicons that cover the full sequence of the virus.

We first examined whether quantitative information on the viral load of the population can be deduced from wastewater sequencing data. Since SARS-CoV-2 enters wastewater essentially via human feces, co-amplification of a virus present in human feces appeared a promising strategy to obtain fecal load normalized SARS-CoV-2 data. We chose PMMoV (pepper mild mottle virus), a virus that is ingested with pepper-containing food. PMMoV is found at high concentrations in human feces and has previously been used for fecal load normalization of wastewater samples [10, 11]. To generate PMMoV normalized data, we included a PMMoV specific primer pair into pool 1 of the ARTIC SARS-CoV-2 primer panel and calculated for each sample the ratio between SARS-CoV-2 and PMMoV reads. Correlation of the SARS-CoV-2/PMMoV ratios with N-gene qPCR data for the individual samples revealed a batch variation between different sampling dates (Figure 2A, inset). We speculated that seasonal variations of pepper consumption or changes in food habits caused by COVID-19 related lockdowns might have led to fluctuations of fecal PMMoV load during the sampling period. To correct for those batch variations we used qPCR data to calculate a correction factor for each sampling date (see Supplementary Table 2 for details). Figure 2A shows that there is a linear relationship between normalized SARS-CoV-2/PMMoV ratios and the number of viral copies derived from RT-qPCR. In this setting, PMMoV allows normalization of samples from the same date while seasonal variations require correction with qPCR data.

**Figure 2:**
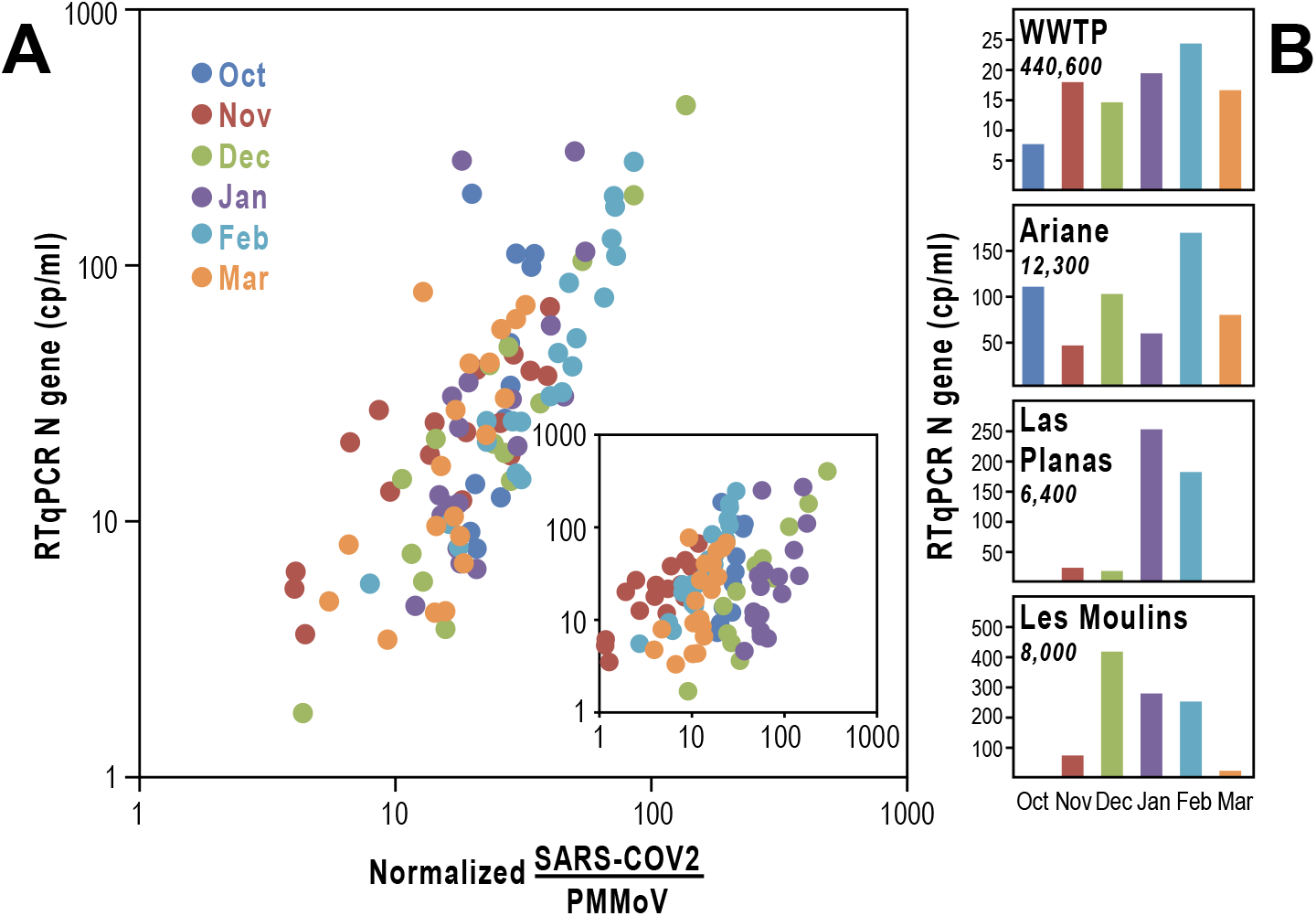
Comparison between RTqPCR and sequencing quantifications. **(A)** Relationship between normalized SARS-CoV-2 / PMMoV ratio and the average RTqPCR signal for the N gene. For each sampling date, a correcting ratio was defined by the weighted average signal between the different sampling points in the wastewater treatment plant (detailed in Supplementary Table 2). The inset shows the relationship between the SARS-CoV-2 / PMMoV ratio and the average RTqPCR signal for N gene before normalization. **(B)** N gene RT-qPCR Cq values were used to assess the concentrations of virus in the WWTP (Haliotis, Nice wastewater treatment plant) and the different neighborhoods. Values are provided in copies of genomes per ml of wastewater (cp/ml). Results are shown for 4 areas. Full results are provided in Supplementary Table 2. The population in each neighborhood is indicated under each name in italic.

The weather conditions and the biological oxygen demand at 5 days (BOD5) of the wastewater remained stable at the different sampling times, as indicated by the stability of the flow rate and of the population equivalent (Supplementary Table 3). The observed variations of virus concentration in wastewater can therefore be interpreted as an appropriate indicator of the SARS-CoV-2 shedding rate by infected communities. Figure 2B indicates for the WWTP “Haliotis” an increase of the virus concentration from October to February followed by a decline in March, which was consistent with other observations [12]. We noticed however different time courses and magnitudes of virus titers in different neighborhoods, possibly due to different modes of circulation of the virus. For instance, a strong peak appeared in “Les Moulins” between December and February, and a single peak was observed in January and February in “Las Planas”. These observations are consistent with the view that a higher population density, a characteristic of these socially deprived districts, is often associated with a higher circulation of the virus.

Two main objectives of our study were also : (1) to define the relative abundance of SARS-CoV-2 lineages and (2) to identify novel SARS-CoV-2 variants in wastewater samples. Available bioinformatics workflows for SARS-CoV-2 sequencing data were not directly suitable for wastewater data since they were designed for the analysis of individual patient data, where each sample is assigned to one single lineage. We therefore designed a dedicated bioinformatics workflow where we first generated PCR primer trimmed, SARS-CoV-2 aligned reads with the ARTIC pipeline. We subsequently counted the mutation frequency at each position of the virus with the iVar software and assigned lineages based on mutational signatures defined in the Pangolin Github repository [13] with a custom pipeline written in R (detailed in the methods section). The associated code is available at https://github.com/ucagenomix/cagablea. A global overview of the mutations found in the samples and the associated lineages is presented in Figure 3.

**Figure 3.**
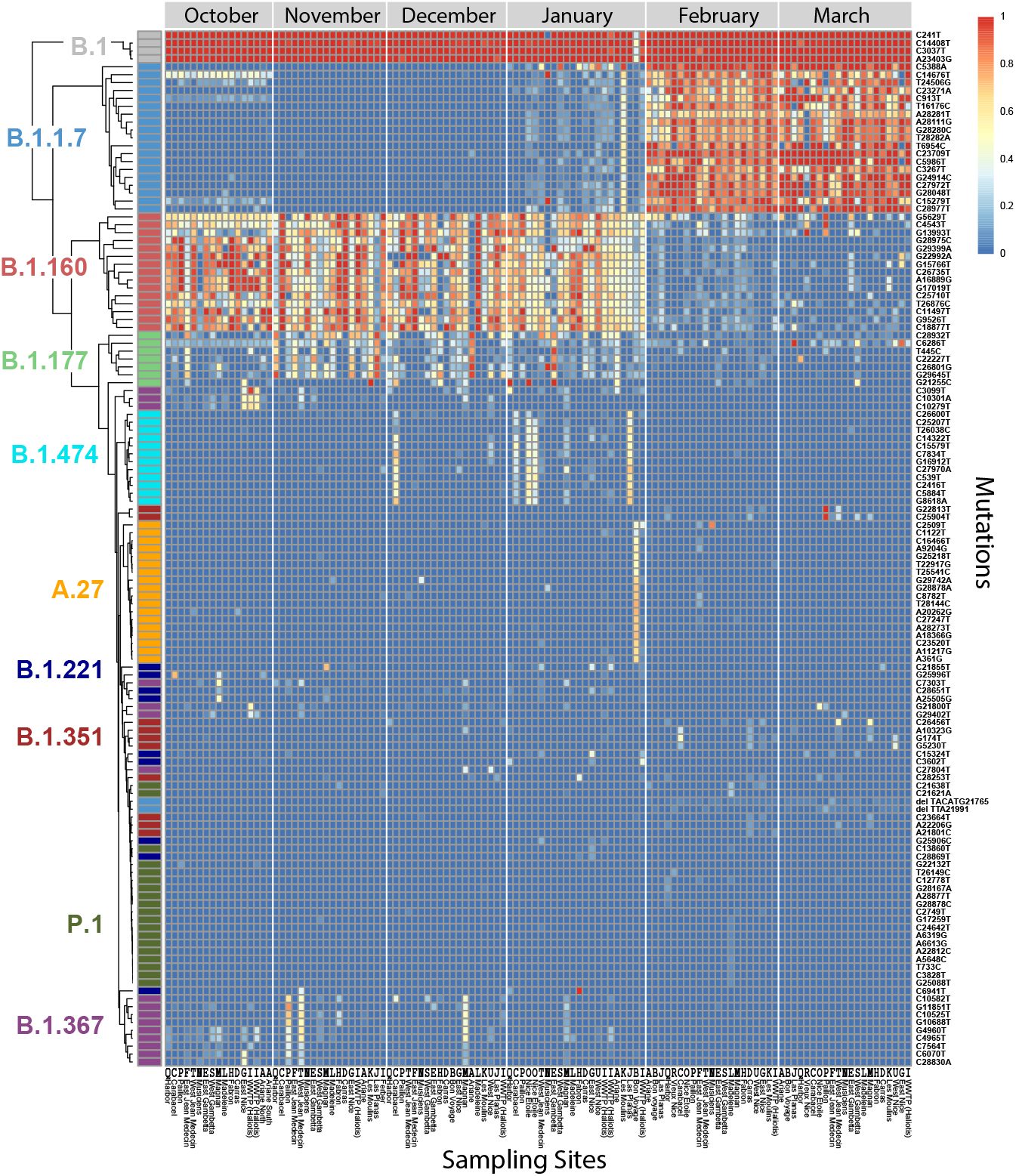
Mutation frequencies for the polymorphisms that define the ten most abundant lineages. Heatmap representation for ten lineages, grouped by dates of collection. For each lineage, the mutations characteristic for the lineage are shown (total: 179 mutations, rows) for each month and sampling site (columns). The color of the boxes indicates the fraction of the reads with the given mutation (blue 0%, red 100%). Reference of the different sampling sites (columns): A, Ariane; B, Bon Voyage; C, Carabacel; D, Carras, E, East Gambetta; F, East Jean Médecin; G, East Nice; H, Fabron; I, WWTP Haliotis; J, Las Planas; K, Les Moulins; L, La Madeleine; M, Magnan; N, Musiciens; O, Nice Etoile; P, Paillon; Q, Harbour; R, Vieux Nice; S, West Gambetta; T, West Jean Médecin; U, West Nice.The same letter code is used in Figures 1 and 4.

The percentage of a lineage was defined as the median of the ratios (number of reads)_variant_ / (number of reads)_total_ for the mutations defining the variant. When mutations that define a lineage are located on amplicons derived from two distinct primer pools (distinct PCR amplifications, Figure 1), the lineage assignment correlated well when the data for each primer pool was analyzed separately. In some cases, a variant was defined by several mutations located on the same amplicon (B.1.351: Figure 3E; B.1.1.7: Supplementary Figure 2A). The search for multiple mutations on the same read allowed us to detect weakly expressed lineages with a high degree of confidence (B.1.351, Figure 3E). The available sequencing information was generally sufficient to ensure robust lineage determination (Figure 3A-C). Nine lineages were enough to cover most of the diversity found in Nice (Figure 3B). From October 2020 to the end of January 2021, the most abundant lineage was B.1.160. Additional lineages (B.1.177, B.1.367, B.1.474, B.1.221, A.27) appeared sporadically in some neighborhoods, but in general, their frequency subsequently decreased rapidly (Figure 3). Most striking was the transient appearance of the A.27 lineage in the “Bon Voyage” neighborhood in January 2021, where it represented half of the virus population (Figure 3 and Figure 4). This lineage had been previously reported in the French overseas department Mayotte, then in Créteil [14] and the detection of this variant in Nice in January may be related to the arrival of travelers from these areas for Christmas. A.27 totally disappeared in February in this neighborhood when B.1.1.7 became prominent. The cumulative fraction of the different lineages shown in Figure 3B was usually around 100%. The missing signal usually corresponded to the B.1 or B.1.1l lineages which were not represented in Figure 3B, because these 2 variants overlap with most of the other observed lineages (Figure 3A).

**Figure 4.**
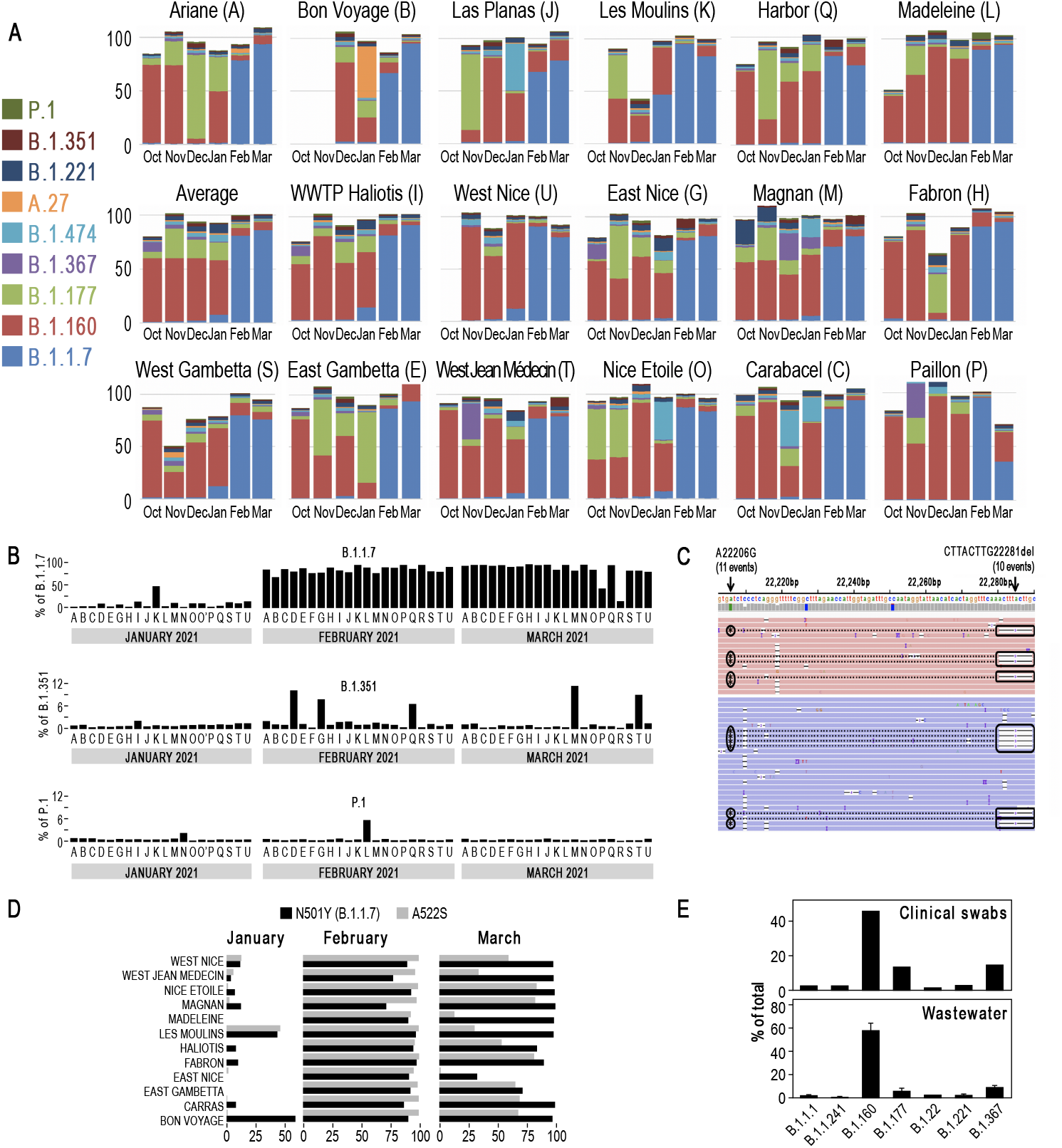
Characterization of SARS-CoV-2 lineages in the 113 samples. **(A)** Barplots illustrating the relative abundance of 9 lineages in the Haliotis WWTP and 17 neighborhoods. **(B)** Barplot showing the fraction of the B.1.1.7, B.1.351 and P.1 lineages in the different Nice neighborhoods. The correspondence between the letter code and the different neighborhoods is provided in Figure 1 and in the legend of Figure 3. Supplementary Figure 3 shows additional barplots for B.1.525 and A.23.1. **(C)** Association of two B.1.351 mutations in the same read. Shown for a sample from “Magnan’’ collected in March 2021. **(D)** Identification of a B.1.1.7 variant, characterized by the presence of an additional A522S (G23126T) mutation in the Spike protein. **(E)** Comparison of the frequencies of different lineages in clinical and wastewater samples both from week 43 (October 2020). Error bars for wastewater data represent SD of the frequencies of the mutations characterizing the variant. See also Supplementary Table 4.

The most striking observation is the appearance of B.1.1.7 in Nice in early 2021. This lineage was found in January 2021 in “Les Moulins” where it represented 50% of the reads (Figure 3, Figure 4A, B). B.1.1.7 was then also detected in “West Nice” and “Haliotis” where it represented 10-20% of the virus population. This is consistent with the fact that these two wastewater networks are collecting sewage from “Les Moulins” (Figure 1A). B.1.1.7 was also detected at lower levels in “Carras”, “Nice Etoile”, “Magnan” and “West Gambetta”. The fraction of this strain then strongly increased in the whole city in February to 81±1% of the total sequences. B.1.1.7 remained predominant in March, representing 86±2% of the viruses circulating in the territory.

Interestingly, the majority of the B.1.1.7 variant that appeared in January in Nice had an additional A522S (G23126T) mutation in the spike protein (Figure 4D). This A522S spike protein mutation had been reported in only 226 out of 27,268 (0.8%) and 207 out of 385,840 (0.05%) GISAID submitted B.1.1.7 sequences for France and the UK, respectively [15]. This variant remained predominant in February and subsequently declined in March (Figure 4D). The initial prevalence of this elsewhere rare B.1.1.7 mutant in Nice suggests that the B.1.1.7 variant was introduced by one or a few carriers and subsequently spread initially in “Les Moulins” and later in the entire city. This variant then became gradually diluted by B.1.1.7 lacking the A522S mutation which is predominant in France. The decline of the A522S mutation in March also suggests that this additional spike mutation does not provide improved fitness when compared to the original B.1.1.7 lineage. This is in line with a recent SARS-CoV-2 / ACE2 interaction modelling study that only found a slight increase of SARS-CoV-2 receptor affinity for the A522S mutation [16].

By contrast, the circulation of other VOCs, such as B.1.351 and P.1, remained much lower (Figure 4B, Supplementary Figure 3A-B). Figure 3 shows that some mutations that are associated with these lineages could be detected at the end of 2020. As we only detected a fraction of the mutations defining those VOCs, we concluded that they were not specific to VOCs at this time. In only a few cases, a consistent signature with enough VOC-specific mutations could be identified. This was for instance the case for “B.1.351” in “Carras”, “East Nice” and “Harbor” in February 2021. This presence was further demonstrated by the detection of two B.1.351 specific mutations on the same reads (Figure 4C). The P.1 lineage represented about 10% of the reads in “La Madeleine” in February 2021, but it was not detected there in March anymore (Fig. 4B).

We finally compared the lineage composition of the October wastewater sample with those of matched clinical samples. The 1,481 swab samples from patients living in Nice that were tested by Biogroup during a period corresponding to our October wastewater sampling (between Oct 19^th^ and Oct 23^rd^, Week 43) were analyzed. There were 1,327 negative and 154 positive SARS-CoV-2 RT-qPCRs. We sequenced 81 PCR positive samples that could be unambiguously assigned to one of the wastewater catchment areas and determined their lineages (Figure 3D, Supplementary Table 4). We found similar percentages for the four most abundant lineages in wastewater and in clinical samples that were collected in the same week in the same area.

In summary, we defined the major lineages of SARS-CoV-2 that circulated from October 2020 to March 2021 in Nice. We were able to identify the switch that occurred between January and February 2021, characterized by a rapid onset of the B.1.1.7 lineage, and a joint decrease of the previously dominant lineages. As of February 2021, B.1.1.7 became the major lineage present in the city, representing more than 80% of all sequences. Our work shows small, localized spikes of the variants of concern B.1.351 and P.1 which did not spread widely into the whole city.

Altogether, our results demonstrate the reliability of wastewater RNA sequencing and the interest of this approach for the monitoring of the emergence and spread of SARS-CoV-2 variants. A limitation of our study is that our analyses were done with RNA from 1 milliliter of wastewater. An increase of the amount of RNA led to RT/PCR inhibition likely due to the co-purification of inhibitors present in wastewater during virus concentration and RNA preparation. Future SARS-CoV-2 concentration techniques and RT/PCR inhibitor removal from the RNA preparations will certainly improve the sensitivity of the measurement, as previously done for detecting poliovirus [17]. This approach was however sufficient in the present study because the concentration of the virus was high at that time in Nice. The results were robust, and we believe that internal controls such as PMMoV will contribute to the development of more quantitative approaches.

In conclusion, our results strongly position RNA sequencing of wastewater samples as a credible tool for analyzing the spread and evolution of SARS-CoV-2. Viral monitoring in wastewater is clearly not limited to the surveillance of the present SARS-CoV-2 pandemic. The surveillance approach that we have set up can easily be transferred to other epidemiological programs, targeting future emerging viruses of concern. As such, it appears as a valuable monitoring tool to assess the effectiveness of sanitary measures aiming to curb virus transmission.

Last, our work illustrates how the situation can vary between different neighborhoods of the same city. The raise of B.1.1.7 in “Les Moulins” in January coincides with a raise of A.27 in “Bon Voyage”, and of B.1.474 in “Las Planas”. The emergence of those particular lineages in three socially deprived neighborhoods occurred immediately after Christmas, a time of very active traveling. This possibly highlights the link existing between socio-economic inequalities between different neighborhoods and the spread of the virus [18]. In this perspective, we believe that wastewater epidemiology could in the future represent a particularly useful instrument to better face similar situations.

## Materials and Methods

### Wastewater sample collection

The characteristics of the different sampling sites are detailed in Figure 1 and Supplementary Table 1, which shows the sizes of the different populations, flow rates and volumes of water for each sampling. Fourteen sampling sites were equipped with automatic sampling devices (Sigma SD900, Hach Company, Loveland, Colorado). The instruments were calibrated to perform twelve samplings per hour for 24 hours. In some cases, especially at the beginning of this study, a manual collection was performed (Supp. Table1). All samples were processed within 24 hours. Sixty milliliters of 0.22µm-filtered wastewater were ultracentrifuged (1 hour, 158,000g, 4°c; Figure 1C). Pellets were resuspended, in 200µl of water, incubated for 5 minutes at room temperature and mixed carefully by pipetting up and down. Viral RNA was extracted using the AllPrep PowerViral DNA/RNA kit (Qiagen, Hilden, Germany, #28000-50) or the RNeasy PowerMicrobiome kit (Qiagen, #26000-50) following the manufacturer’s instructions. RNA was eluted in 100µl of RNase free water and stored at -80°C. All wastewater and clinical samples were handled in a biosafety level 2 laboratory under a biological safety cabinet using adapted personal protective and respiratory equipment.

### Clinical samples

A total of 1409 nasopharyngeal swabs was collected by Biogroup (Montauroux, France) during week 43 (2021) in the area of Nice (France). This represents the full collection of samples that was processed during this week in this area by Biogroup. Out of these 1409 samples, 172 were positive by RTqPCR. We were able to generate SARS-CoV-2 sequencing data for 81 of them. The use of patient samples complied with French legislation. During the health emergency SARS-CoV-2 related patient data and laboratory results in France are recorded in the SARS-Cov2 Screening Information System SI-DEP («Système d’Informations de DEPistage», French decree No. 2020-551 of May 12, 2020, Article 11 of Law No. 2020-546 of May 11, 2020 extending the state of health emergency). In the context of this law, patients can not object to the use of their SARS-CoV-2 related data for health surveys and epidemiological monitoring. Under this legislation, authorization of the use of patient data for research was granted unless patients actively express their opposition.

### RNA extraction

Viral RNA from 81 nasopharyngeal swabs were extracted using the QIAamp Viral RNA minikit (Qiagen, #52904) according to manufacturer instructions. Samples RNA were eluted with 60µl of AVE buffer from the kit (0.04% of sodium azide in nuclease free water) and stored at -80°C before further analysis.

### Gene N1 RTqPCR

For SARS-CoV-2 RNA quantification by RT-qPCR, the N1 region of the nucleocapsid gene was targeted using the primers and probes as well as the PCR conditions described in the CDC diagnostic panel (2020). RT-qPCR quantifications were performed using the qScript XLT One-Step RT-qPCR ToughMix (Quantabio) on CFX96 touch Real-Time PCR Detection System. The N1 probe was labelled at the 5’-end with the FAM reporter and with the BHQ-1 quencher at the 3’-end. For each sample, RT-qPCR analysis was systematically performed in duplicate of which the mean is reported in the study.

Prior to the RT-qPCR, extracted RNAs were diluted with RNase free water to bring the initial RNA amount back in a range of [50-150 ng] RNA quantity per reaction. When the RNA concentration was below 50 ng, the sample was by default diluted by a factor of 2, in order to dilute matrix-related inhibitors. An inhibition control of the RTqPCR reaction was systematically performed for each extracted sample. This control consisted of a dosed addition (∼4000 copies/reaction) of the positive control EDX SARS-CoV-2 (Exact Diagnostic, supplied by Bio Rad) for each dilution analyzed. The amplification was considered as free of inhibition with the Cq 24 with a tolerance of 2 Cq. A standard curve, from 10^6^ copies/μL to 1 copy/μL, was generated for every analysed plate using a control plasmid, N-nCoV-control-Plasmid (Eurofins). Standard curves were performed by serial 10 fold dilution with 1X Tris - EDTA. A negative control was analyzed on each plate, using SARS-CoV-2 Negative Run Control (Exact Diagnostic, supplied by Bio Rad). This control is formulated in a synthetic matrix and contains 75,000 copies/mL of human genomic DNA.

### SARS-COV-2 genome amplification and sequencing

Library preparation was performed using the nCoV-2019 sequencing protocol v3 LoCost [19] with a few modifications. For reverse-transcription, 8 µl of RNA and 2µl of LunaScript (NEB) was mixed and incubated at 25°C for 5 minutes followed by an incubation at 55°C for 10 minutes before holding at 4°C. For cDNA amplification we used the SARS-CoV-2-specific version 3 primer set designed by the ARTIC Network (total 218 primers in two pools, purchased from Integrated DNA Technologies) and the PMMoV specific primers (5-’GGCATGTCTCTATGCTCCGAGGC-3’, 5-’CCCACGGGTGTAGGCGTCAG-3’). One multiplex-polymerase chain reaction was performed for each of both SARS-CoV-2 primer pools by combining 12,5 μl of Q5 Hot Start DNA Master Mix (NEB) and 2.5μl of the reverse transcribed cDNA with either 2μl Nuclease-free water (Qiagen) and 4.00μl of 10μM “Primer Pool #1” and 4.0 µl of 100 nM PMMoV primers or, 6µl Nuclease-free water and 4 uL 10μM “Primer Pool #2”.

The mix was incubated at 98°C for 30 seconds followed by 38 cycles at 98°C for 15 seconds and 65°C for 5 minutes before holding at 4°C.

Pool #1 and Pool #2 were cleaned up separately by adding 1 vol. SPRI select (Beckman Coulter). Samples were eluted in 10μl Nuclease-free water and quantified using a Qubit® High Sensitivity Kit (ThermoFisher) as per manufacturer’s instructions to ensure each amplicon into end-prep preparation.

The end-prep reaction was prepared as follows: 100ng each of Pool #1 and Pool #2 PCR product were mixed with 1.2 μl Ultra II end prep buffer, 0.5 μl Ultra II end prep enzyme (New England Biolabs) and water to a final volume of 10 μl.. The reaction was incubated at room temperature for 15 min and 65 °C for 15 min, followed by a hold at 4 °C for at least 1 min. DNA was purified with 1 vol. SPRI select (Beckman Coulter), eluted in 8.5µL Nuclease-free water (Qiagen) and quantified using a Qubit® High Sensitivity Kit (ThermoFisher) as per manufacturer’s instructions.

Addition of sample barcodes: 7.5 μl end-prepped samples, 2,5 μl barcode (ONT,native barcoding EXP-NBD196)), 10 μl NEBNext Quick T4 Ligase were mixed and incubated at room temperature for 20 min and 65 °C for 15 min, followed by a hold at 4 °C for at least 1 min. The ligation reaction was subsequently cleaned up with 0.4 vol. SPRI select (Beckman Coulter), eluted with 31 µl Nuclease-free water (Qiagen) and quantified (Qubit® High Sensitivity Kit).

Oxford Nanopore sequencing adapter ligation was performed with the Oxford Nanopore native barcoding kit and the Quick ligation kit (New England Biolabs). 600 ng of barcoded cDNA in 30 μl, 5 μl Adapter Mix II (ONT), 10 μl 5X NEBNext Quick Ligation Reaction Buffer (NEB), 5 μl Quick T4 DNA Ligase (NEB). The mix was incubated at room temperature for 20 min bound to 1 vol. (50 μl) SPRI select,, washed twice with 250 μl SFB (Oxford Nanopore ligation sequencing kit SQK-LSK110). The library was eluted in 15 μl of EB buffer (ONT) and quantified.

150 ng of the library was loaded onto a PromethION flow cell. An average of 241,000 ± 20,000 SARS-CoV-2 reads was generated for each sample (∼400 base pair, average read length).

### Bioinformatics

A dedicated bioinformatic pipeline was adapted from ARTIC, Pangolin and Ivar to identify the main circulating lineages. PCR primers were trimmed and reads were mapped using the ARTIC pipeline [9]. The iVar tool [20] was used to generate for each sample a table with mutation frequency data and p-Values for altered positions of SARS-CoV-2 from the BAM files. iVar was run with a minimum base quality filter of 20 using the reference genome of SARS-CoV-2 (MN908947) and the feature file Sars_cov_2.ASM985889v3.101.gff3 from NCBI.

The tables generated by iVar were filtered for positions that were covered by at least 100 reads and for alterations with an adj p-value (Fisher’s test) below 10^−8^.

A custom R-script was implemented to fix a known issue with the iVar output, the incorrect assignment of deletion frequencies and to code the mutations events in amino acid format.

A database with the mutations associated with known lineages was built from the publicly available data [21].

Every mutation event in the sample was associated with mutations described in the database and matched to one or several lineages. Supplementary Figures 4BC indicate that the percentage of B.1.351 (B) and P.1 (C) that were obtained with two independent pools of ARTIC primers provided similar percentages of VOCs.

## Data Availability

All relevant data, custom software, R figures and analysis scripts are available on Github (https://github.com/ucagenomix/cagablea).

https://github.com/ucagenomix

## Data and code availability

All relevant data are accessible through a dedicated webpage. They have also been deposited in Gene Expression Omnibus under accession number GSEXXX (https://www.ncbi.nlm.nih.gov/geo/query/acc.cgi?acc=GSE). All custom software, R figures and analysis scripts are available on Github (https://github.com/ucagenomix/cagablea).

## Acknowledgements + Fundings

PB’s group is supported by FRM (DEQ20180339158). The present work was possible thanks to the support of the National Infrastructure France Génomique (Commissariat aux Grands Investissements, ANR-10-INBS-09-03, ANR-10-INBS-09-02). BM is supported by Plan Cancer 2018 « ARN non-codants en cancérologie: du fondamental au translationnel » (Inserm number 18CN045) and Cancéropole PACA. We are particularly grateful to Delphine Zapalski, Hugues Decobecq, Jean Lafay and Jean-Marc Campeggio, at the Wastewater Department at Nice Métropole Côte d’Azur for their excellent support throughout this project. We would like to thank the GISAID Initiative and are grateful to all of the data contributors, i.e.the Authors, the Originating laboratories responsible for obtaining the specimens, and the Submitting laboratories for generating the genetic sequence and metadata and sharing via the GISAID Initiative, on which this research is based.

---

**Supplementary Figure 1.**
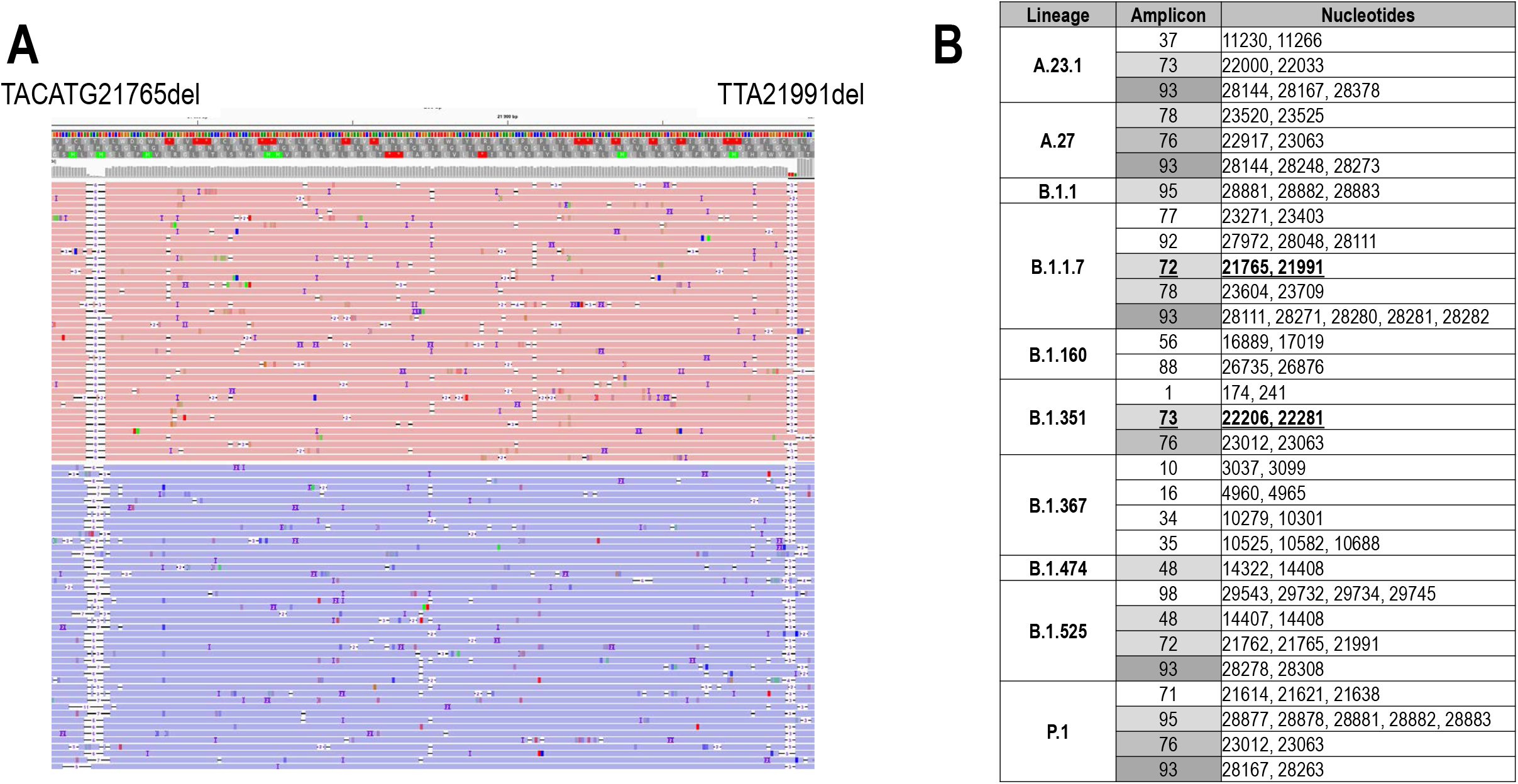
**(A)** High specificity of detection of specific lineages by the presence of 2 mutations in a same read. This is illustrated in Supplementary Figure 2A by the double-deletion in TACATG21765del and in TTA21991del that is associated with B.1.1.7 or B.1.525 lineages. See also Figure 3D. **(B)** Table illustrating amplicons that contain several mutational events for a same lineage. Amplicon 93, for instance, contains mutations found in half of these lineages. Amplicons 72 and 73, shown in Supplementary Figure 2A and Figure 3D, respectively, are underlined.

**Supplementary Figure 2.**
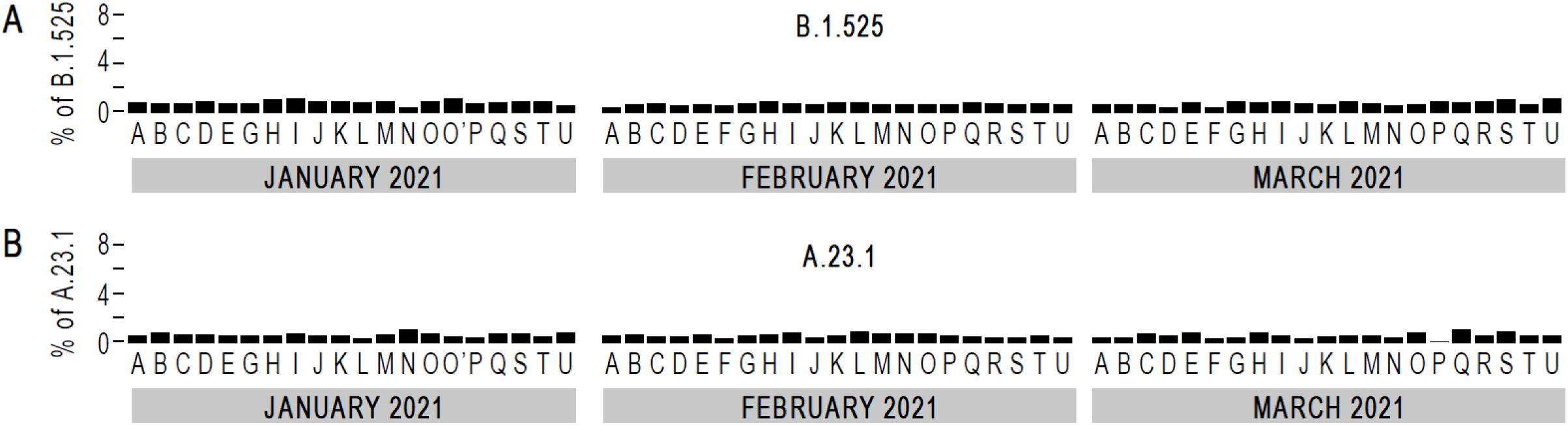
**(A,B)** Barplot representation for the B.1.525 and A.23.1 lineages (A and B, respectively), showing that these lineages were not detected in Nice between January and March 2021. Correspondence between letter code and the different neighborhoods is as follows: “Ariane” (A), “Bon Voyage” (B), “Carabacel” (C), “East Gambetta” (D), “East Jean Medecin” (E), “East Nice” (F), “Fabron” (G), “WWTP-Haliotis” (H), “Las Planas” (I), “Les Moulins” (J), “Madeleine” (K), “Magnan” (L), “Musiciens” (M), “Nice Etoile” (N), “Paillon” (O), “Port” (P), “Vieux Nice” (Q), “West Gambetta” (R), “West Jean Medecin” (S), “West Nice” (T). The same letter code was also used in Figure 3C.

**Supplementary Figure 3.**
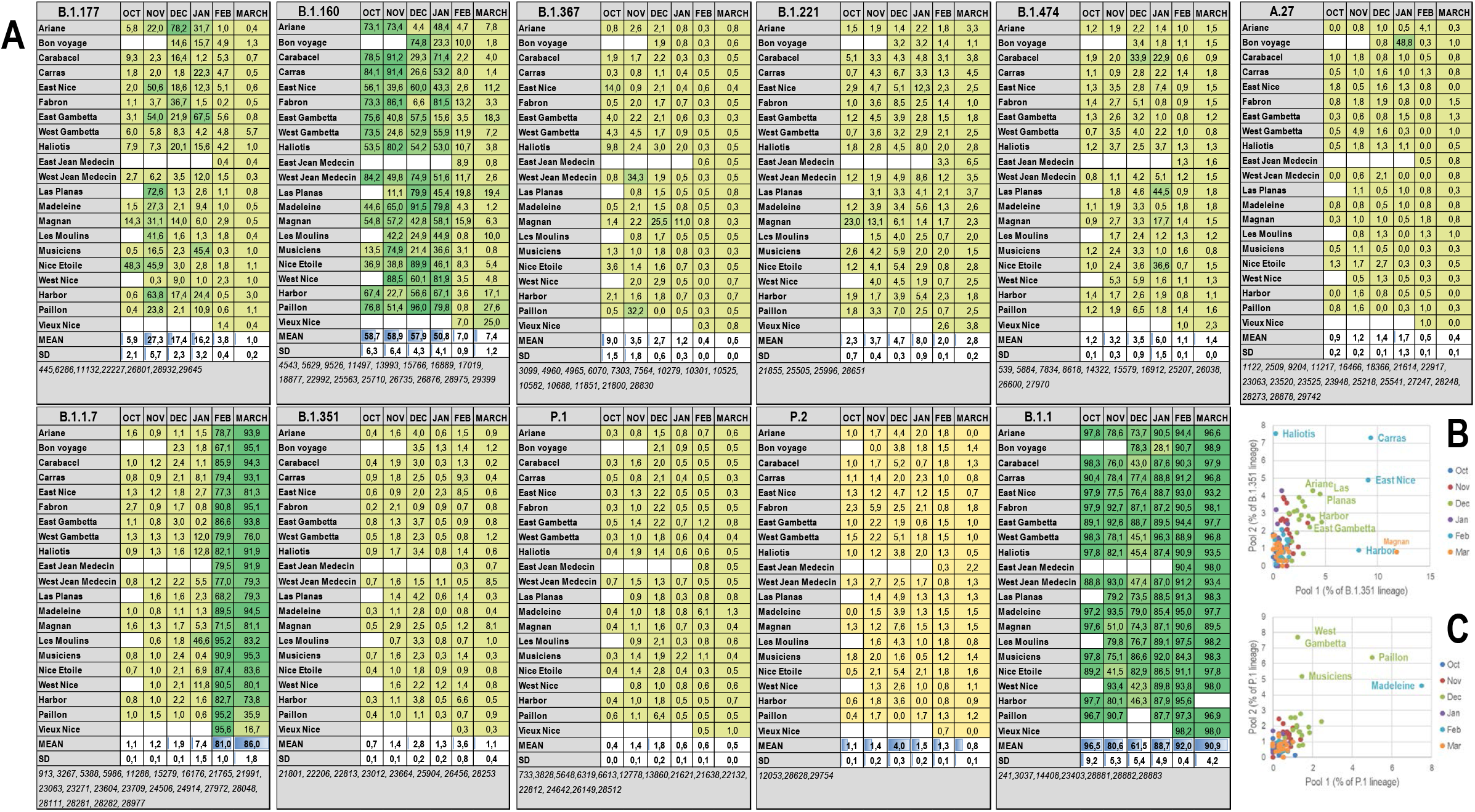
(A) Distribution tables of the main lineages detected in the Nice wastewater samples between Oct. 2020 and March 2021. The neighborhoods are those defined in Figure 1A. The position of the different nucleotides used to assess each lineage is indicated on the last line of each table. **(B, C)** comparison of the ratio MUT/TOT for nucleotides identified in Pool1 and Pool2 sequences. Results are shown for B.1.351 (B) and P.1 (C).

## References

1. Pettit, S.D., et al., ‘All In’: a pragmatic framework for COVID-19 testing and action on a global scale. EMBO Mol Med, 2020. 12(6): p. e12634.

2. Zheng, S., et al., Viral load dynamics and disease severity in patients infected with SARS-CoV-2 in Zhejiang province, China, January-March 2020: retrospective cohort study. BMJ, 2020. 369: p. m1443.

3. Medema, G., et al., Implementation of environmental surveillance for SARS-CoV-2 virus to support public health decisions: Opportunities and challenges. Curr Opin Environ Sci Health, 2020. 17: p. 49–71.

4. Peccia, J., et al., Measurement of SARS-CoV-2 RNA in wastewater tracks community infection dynamics. Nat Biotechnol, 2020. 38(10): p. 1164–1167.

5. La Rosa, G., et al., First detection of SARS-CoV-2 in untreated wastewaters in Italy. Sci Total Environ, 2020. 736: p. 139652.

6. Ahmed, W., et al., First confirmed detection of SARS-CoV-2 in untreated wastewater in Australia: A proof of concept for the wastewater surveillance of COVID-19 in the community. Sci Total Environ, 2020. 728: p. 138764.

7. Wu, F., et al., SARS-CoV-2 Titers in Wastewater Are Higher than Expected from Clinically Confirmed Cases. mSystems, 2020. 5(4).

8. Wurtzer, S., et al., Evaluation of lockdown effect on SARS-CoV-2 dynamics through viral genome quantification in waste water, Greater Paris, France, 5 March to 23 April 2020. Euro Surveill, 2020. 25(50).

9. https://artic.network/ncov-2019.

10. Bivins, A., et al., Wastewater-Based Epidemiology: Global Collaborative to Maximize Contributions in the Fight Against COVID-19. Environ Sci Technol, 2020. 54(13): p. 7754–7757.

11. Zhang, T., et al., RNA viral community in human feces: prevalence of plant pathogenic viruses. PLoS Biol, 2006. 4(1): p. e3.

12. https://www.reseau-obepine.fr/nice/.

13. Rambaut, A., et al., A dynamic nomenclature proposal for SARS-CoV-2 lineages to assist genomic epidemiology. Nat Microbiol, 2020. 5(11): p. 1403–1407.

14. Fourati, S., et al., Novel SARS-CoV-2 Variant Derived from Clade 19B, France. Emerg Infect Dis, 2021. 27(5): p. 1540–1543.

15. Elbe, S. and G. Buckland-Merrett, Data, disease and diplomacy: GISAID’s innovative contribution to global health. Glob Chall, 2017. 1(1): p. 33–46.

16. Wang, R., et al., Vaccine-escape and fast-growing mutations in the United Kingdom, the United States, Singapore, Spain, India, and other COVID-19-devastated countries. Genomics, 2021. 113(4): p. 2158–2170.

17. Berchenko, Y., et al., Estimation of polio infection prevalence from environmental surveillance data. Sci Transl Med, 2017. 9(383).

18. Horton, R., Offline: COVID-19 is not a pandemic. Lancet, 2020. 396(10255): p. 874.

19. https://www.protocols.io/view/ncov-2019-sequencing-protocol-v3-locost-bh42j8ye.

20. https://github.com/andersen-lab/ivar.

21. https://covidcg.org/.

